# Beyond the new normal: assessing the feasibility of vaccine-based elimination of SARS-CoV-2

**DOI:** 10.1101/2021.01.27.20240309

**Authors:** Madison Stoddard, Sharanya Sarkar, Ryan P. Nolan, Douglas E. White, Laura F. White, Natasha S. Hochberg, Arijit Chakravarty

## Abstract

As the COVID-19 pandemic drags into its second year, there is hope on the horizon, in the form of SARS-CoV-2 vaccines which promise disease elimination and a return to pre-pandemic normalcy. In this study we critically examine the basis for that hope, using an epidemiological modeling framework to establish the link between vaccine characteristics and effectiveness in bringing an end to this unprecedented public health crisis. Our findings suggest that vaccines that do not prevent infection will allow extensive endemic SARS-CoV-2 spread upon a return to pre-pandemic social and economic conditions. Vaccines that only reduce symptomatic COVID-19 or mortality will fail to mitigate serious COVID-19 mortality risks, particularly in the over-65 population, likely resulting in hundreds of thousands of US deaths on a yearly basis. Our modeling points to the possibility of complete SARS-CoV-2 elimination with high population-level compliance and a vaccine that is highly effective at reducing SARS-CoV-2 infection. Notably, vaccine-mediated reduction of transmission is critical for elimination, and in order for partially-effective vaccines to play a positive role in SARS-CoV-2 elimination, other stackable (complementary) interventions must be deployed simultaneously.

## Introduction

The ongoing COVID-19 pandemic, caused by the novel coronavirus SARS-CoV-2, is the health crisis of our lifetimes, placing a burden on global health and economic well-being that would have been inconceivable to us at the start of 2020. The biomedical community has responded with unprecedented speed, and several vaccines are now being rolled out worldwide. The experiences of 2020 have shown that the high basic reproduction number (R_0_) and capacity for presymptomatic spread make SARS-CoV-2 difficult to control once it is established within a population [1,2]. Thus, if we are ever to return to a pre-pandemic normal, a focus on SARS-CoV-2 elimination will be critical, and vaccines are expected to play a crucial role in this endeavor.

For SARS-CoV-2, an efficacious vaccine might prevent colonization of the nasal cavity (referred to in this work as infection), symptomatic disease, severe disease, mortality or transmission [3]. In the clinical trials for the front-runner vaccines (AstraZeneca/Oxford, Moderna and Pfizer/BioNTech), the primary endpoint is symptom-gated efficacy, where cases are counted as patients presenting with symptoms of COVID-19 and SARS-CoV-2 infection confirmed by nucleic acid amplification test.

The characteristics of SARS-CoV-2 raise a number of risks for vaccine-based elimination strategies: the waning nature of both natural [4] and vaccine-mediated immunity [5], the potential for vaccinated individuals to transmit the disease [6], the age-dependence of both disease severity [7] as well as (potentially) vaccinal immunity [8,9] and the potential for the emergence of resistance [10,11].

Waning natural immunity, originally proposed as a potential risk for SARS-CoV-2 based on the behavior of other coronaviruses [12,13] has now been demonstrated for cellular [14] as well as humoral [4,15] immunity in response to COVID-19. Numerous cases of SARS-CoV-2 reinfection have been proven using direct molecular techniques [16,17]. The relative infrequency of such events has been shown to be only a consequence of epidemiological dynamics– in a situation where the uninfected population is larger than the previously-infected population, reinfections will make up a small proportion of new infections [18], but the rate-limiting step is the percentage of previously-infected individuals and not the duration of immunological protection. The potential for relatively short-term immunity complicates disease control by limiting the duration of natural herd immunity and providing the virus with a foothold even in populations that have previously experienced a high attack rate [19]. If vaccinal immunity were to also wane over similar timeframes, the logistical burden of a vaccine-based elimination strategy for this disease may become insurmountable.

Asymptomatic transmission by vaccinated but colonized individuals poses another risk for vaccine-based elimination of SARS-CoV-2. Asymptomatic SARS-CoV-2 infections have been demonstrated to present with viral loads similar to symptomatic infections [20]. Contact tracing and modeling studies suggest presymptomatic and asymptomatic patients are responsible for approximately 50% of all COVID-19 transmission events [21,22]. While prevention of infection (sterilizing immunity) is the norm for many vaccines, there are a number of vaccines (such as the inactivated polio vaccine [23] and the polyvalent pneumococcal vaccine [24]) that merely decrease the potential for onward transmission from colonized but vaccinated individuals. Preclinical studies for a number of COVID-19 vaccine candidates have demonstrated viral infection of the nasal cavity in vaccinated primates [25,26]. For the AstraZeneca vaccine, estimates for the reduction in frequency of asymptomatic COVID-19 cases in the vaccinated arm (measured by seroconversion) ranged from 4% to 59% (95% confidence interval 1 to 83%) depending on dose schedule [6,27]. The Pfizer vaccine showed a 9% reduction in seroconversion for the vaccinated arm [8], and vaccinated asymptomatic carriers were observed in the Moderna clinical trials as well [28]. To the extent that vaccinated patients have their nasal cavities colonized by SARS-CoV-2, they will very likely present as being asymptomatic, and be able to transmit the disease (albeit possibly at a lower frequency [29]), making it further challenging to achieve herd immunity.

The age structure of morbidity and mortality of COVID-19 is further unhelpful in the context of the age-dependence of benefits derived from vaccination. For every two decades of age, the infection fatality rate (IFR) for COVID-19 rises by about one order of magnitude, ranging from 0.013% at 25 years to about 8% at 85 years [7]. Age-dependent reductions in vaccine efficacy have been demonstrated for a number of other diseases [24,30–32]. For some of the SARS-CoV-2 vaccine candidates, peak antibody titers in the over-65 population appear to be lower relative to younger vaccinees [9]. Subgroup analyses of vaccine efficacy by age group have not yielded definitive results so far, primarily due to sample size and study design issues. (For example, the 95% confidence interval for vaccine efficacy against symptomatic COVID-19 reported by Pfizer is 66.7% to 99.9% for the over-65 age group [8], and the subgroup analysis for the AstraZeneca/Oxford vaccine was not performed due to methodological issues with the trial design resulting in age being confounded with dose schedule [6]).

Widespread, endemic SARS-CoV-2 infection within the population will also create a fertile breeding ground for immune evasion mutants. Viral genomic data collected over the course of the pandemic shows substantial sequence variation for the viral spike protein [33], as well as other surface proteins [34]. Notably, the SARS-CoV-2 genome does not currently appear to be under strong positive selection pressure as a result of the immune response [35,36], consistent with the time lag of about 10 days between peak levels of viral load (and transmissibility [37] and the peak of the host antibody response [38]. The natural immune response is narrowly targeted, with the epitopes for RBD-targeting neutralizing antibodies overlapping one another substantially [39,40]. On the other hand, numerous putative SARS-CoV-2 immune escape mutants with ACE2 affinities comparable to wild type have been demonstrated to be infection-competent *in vitro*, suggesting a low fitness cost for the virus in evading the immune response [41–43]. Evolutionary modeling conducted by us suggests that SARS-CoV-2 immune evasion mutants with one or two mildly deleterious mutations likely pre-exist in high numbers due to neutral genetic variation, and resistance to vaccines targeting multiple epitopes on the spike protein may occur rapidly under certain conditions [10]. Consistent with these predictions, recent reports have noted the emergence of virus strains (e.g. the South African 501Y.V2 variant) that are capable of complete immune escape from therapeutically relevant monoclonal antibodies, as well as convalescent plasma [44].

Taken together, the risks described above represent a set of real-world constraints for any SARS-CoV-2 elimination strategy that relies primarily on vaccines to bring us back to the ‘old normal’. In this study, we use an epidemiological modeling framework to explore the impact of waning natural immunity, unknown vaccine efficacy against infection and transmission, vaccine refusal, and uncertainty in vaccine efficacy against mortality in individuals older than 65 on the feasibility of SARS-CoV-2 elimination using vaccines. We also estimate the mortality burden of COVID-19 under the first wave of vaccines. We further extend this work to examine practically useful strategies for vaccine-based elimination of SARS-CoV-2.

## Methods

### SEIRS Model

To evaluate strategies for achieving elimination of SARS-CoV-2 and to predict the impact of existing vaccines, we built an SEIRS (susceptible-exposed-infectious-recovered-susceptible) model to account for disease spread, waning natural immunity, and the deployment of a vaccine in some or all of the population. The model has two parallel sets of SEIR compartments representing the vaccinated and unvaccinated populations. Although we recognize that vaccine coverage may be geographically heterogeneous, for the purposes of this model we assume all compartments are well-mixed, meaning that vaccinated and unvaccinated individuals are in contact and can infect each other. The vaccine may confer a benefit at any of multiple stages: infection, transmission, and disease progression. Thus, the vaccinated population may have a reduced susceptibility to infection (“reduction in risk of infection”), a reduced transmissibility upon infection (“reduction in risk of transmission”), or a reduced likelihood of downstream negative health outcomes upon infection (“reduction in risk of symptoms”, “reduction in risk of mortality”). We note that risk of infection and risk of transmission are the determinants of SARS-CoV-2 infection risk and spread, while downstream risks of symptomatic COVID-19 or mortality impact the outcomes of the infected population. Vaccinal immunity is assumed to be life-long or continually boosted such that the degree of vaccinal immunity does not wane or vary in vaccinated individuals. This represents the best-case scenario for vaccine effectiveness; the feasibility of frequent repeat vaccinations and constraints on the vaccine deployment timeline are outside the scope of this paper. Additionally, vaccinated individuals who become infected (I_v_) are assumed to develop natural immunity that provides transient protection from reinfection. The ordinary differential equations (ODE)-based SEIRS model is summarized by equations 1-8

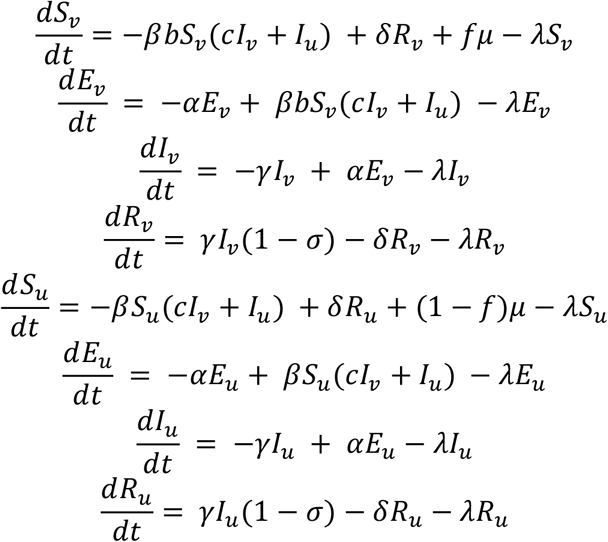

Where *S* represents the susceptible population, *E* represents the exposed population, *I* represents the infectious population, and *R* represents the recovered population. Subscripts *v* and *u* delineate the vaccinated and unvaccinated subpopulations, respectively. Model parameters are summarized in Table 1.

**Table 1:** Model parameters for SEIRS model.

| Parameter | Symbol | Value | Source |
| --- | --- | --- | --- |
| Latency period | $1/\alpha$ | 3 days | [45] |
| Reproductive number | $R_0$ | 5.7 individuals | [46] |
| Infectious period | $1/\gamma$ | 10 days | [47] |
| Duration of natural immunity | $1/\delta$ | 18 months | [48] |
| Infection fatality rate (IFR) | $\sigma$ | 0.68% | [49] |
| Population birth rate | $\mu$ | 1% annually | [50] |
| Population death rate | $\lambda$ | 0.9% annually | [51] |
| Fraction compliant | $f$ | Variable | |
| Vaccine reduction in risk of infection | $1-b$ | Variable | |
| Vaccine reduction in risk of transmission | $1-c$ | Variable | |

The contact rate *β* is derived from the relationship between the intrinsic reproductive number *R*_*0*_, the contact rate, and the infectious period

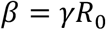

According to the CDC, the *R*_*0*_ for SARS-CoV-2 under pre-pandemic social and economic conditions is estimated to be 5.7 based on data from China in early 2020. Although the *R*_*0*_ is commonly held to be in the range of 2-3 based on early estimates, the CDC’s study provides an updated estimate based on improved methods designed to address stochasticity in early outbreak dynamics and drawn from a larger set of case reports to address the low numbers of observations that early studies were reliant on [46]. For the purpose of this study, an *R*_*0*_ of 5.7 is used to represent epidemiological conditions under a theoretical return to pre-pandemic activity. In the supplement, the simulations are re-implemented with an *R*_*0*_ of 3.32, in keeping with the results reported by a different systematic meta-analysis of the range of *R*_*0*_s reported in the early stages of the pandemic [52].

In the long-term, the SEIRS system is bistable with two steady-state equilibria: either “elimination,” a condition under which yearly SARS-CoV-2 infection rates approach zero, or “endemic disease,” a scenario in which SARS-CoV-2 infection and transmission persist at a constant rate at steady-state determined by the effective population contact rate [53]. In this analysis, we further modified the model to explore the tendency toward elimination or endemic disease under a variety of scenarios.

### Model adaptation: Vaccinated asymptomatic carriers

To address the possibility of a vaccine that prevents COVID-19 symptoms or mortality but not nasopharyngeal SARS-CoV-2 infection, we adapted the model to treat vaccinated individuals as potential asymptomatic carriers. In this case, we explore the impact of vaccine’s efficacy against mortality or symptomatic COVID-19. We assume that the vaccine does not prevent infection but reduces transmission by reducing the incidence of symptoms. We assume that asymptomatic individuals have a 42% reduced transmission rate *c* relative to symptomatic individuals [29]; in the supplement, we explore the impact of this assumption by running the simulations with no reduction in transmission. For these simulations, we track the number of yearly US symptomatic cases and the expected yearly death toll. In the unvaccinated population, asymptomatic cases are expected to account for 20% of total cases [54].

### Model adaptation: Age-variant vaccine efficacy

Unfortunately, vaccines commonly are less effective in older individuals [24,30–32], and older individuals are significantly more susceptible to fatal COVID-19 outcomes [7]. For some candidate SARS-CoV-2 vaccines, antibody titers peak lower [55] and decline more rapidly [5] in the over-65 population. Current clinical trials show a high degree of uncertainty for vaccine efficacy in older volunteers. For example, in the clinical data for Moderna’s vaccine, efficacy against symptomatic COVID-19 is 95.6% in the 18-64 year-old age group (95% CI 90.6% -97.9%). In the over-65 group, vaccine efficacy is reported to be 86.4% with a much wider confidence interval (95% CI 61.4% – 95.5%) [56]. To understand the impact of lower vaccine efficacy in the elderly population on mortality, we conducted epidemiological modeling to assess the steady-state mortality burden, using a standard SEIRS model. Specifically, we simulated a scenario where vaccine efficacy against mortality in the under-65 age group was fixed at 95%, and vaccine protection for the over-65 age group was varied across a range. We considered the over-65 age group to represent 16.4% of the US population, consistent with current demographic estimates [57]. We calculated a population age structure-adjusted IFR of 3.34% among over-65 Americans and an IFR of 0.16% among under-65 Americans based on age-dependent US IFRs [7].

### Model adaptation: Stacked interventions

We also modified the model to simulate disease dynamics in a scenario where two complementary biomedical or nonpharmaceutical interventions are implemented. In this model, there are four sets of SEIR compartments, representing the four permutations of participation and nonparticipation with the two interventions. The choice to participate in each intervention is assumed to be independent: thus, the fraction of the population participating in both interventions is f_1_*f_2_. The population is assumed to be well-mixed with no strategy-switching during the modeled time period. In these scenarios, we track the total number of US infections on a yearly basis at steady-state.

## Results

### Vaccines that do not prevent infection (and hence transmission) allow extensive endemic SARS-CoV-2 spread

If vaccines prevent symptom onset but do not prevent infection, SARS-CoV-2 will become endemic in the population, reaching a steady-state yearly number of cases determined by the duration of natural immunity and the contact rate in the population, which determines R_0_. Table 2 provides model-predicted yearly infections in the United States after extensive vaccination (90% or more of the population) for a set of realistic combinations of R_0_ and durations of natural immunity. Yearly SARS-CoV-2 infections are expected to number in the hundreds of millions in the US.

### A vaccine that only reduces symptomatic COVID-19 or mortality is a limited solution

The elimination of COVID-19 is virtually impossible without eliminating SARS-CoV-2 (Fig. 1, 2). The high expected yearly infection rate for SARS-CoV-2 implies a high disease and mortality burden in the absence of intervention: 159 million infections resulting in 127 million symptomatic cases and 1.1 million deaths could be expected. A vaccine effective against symptomatic disease and mortality could mitigate such a catastrophic scenario but cannot achieve SARS-CoV-2 elimination or provide communal benefit via “herd immunity.”

**Figure 1.**
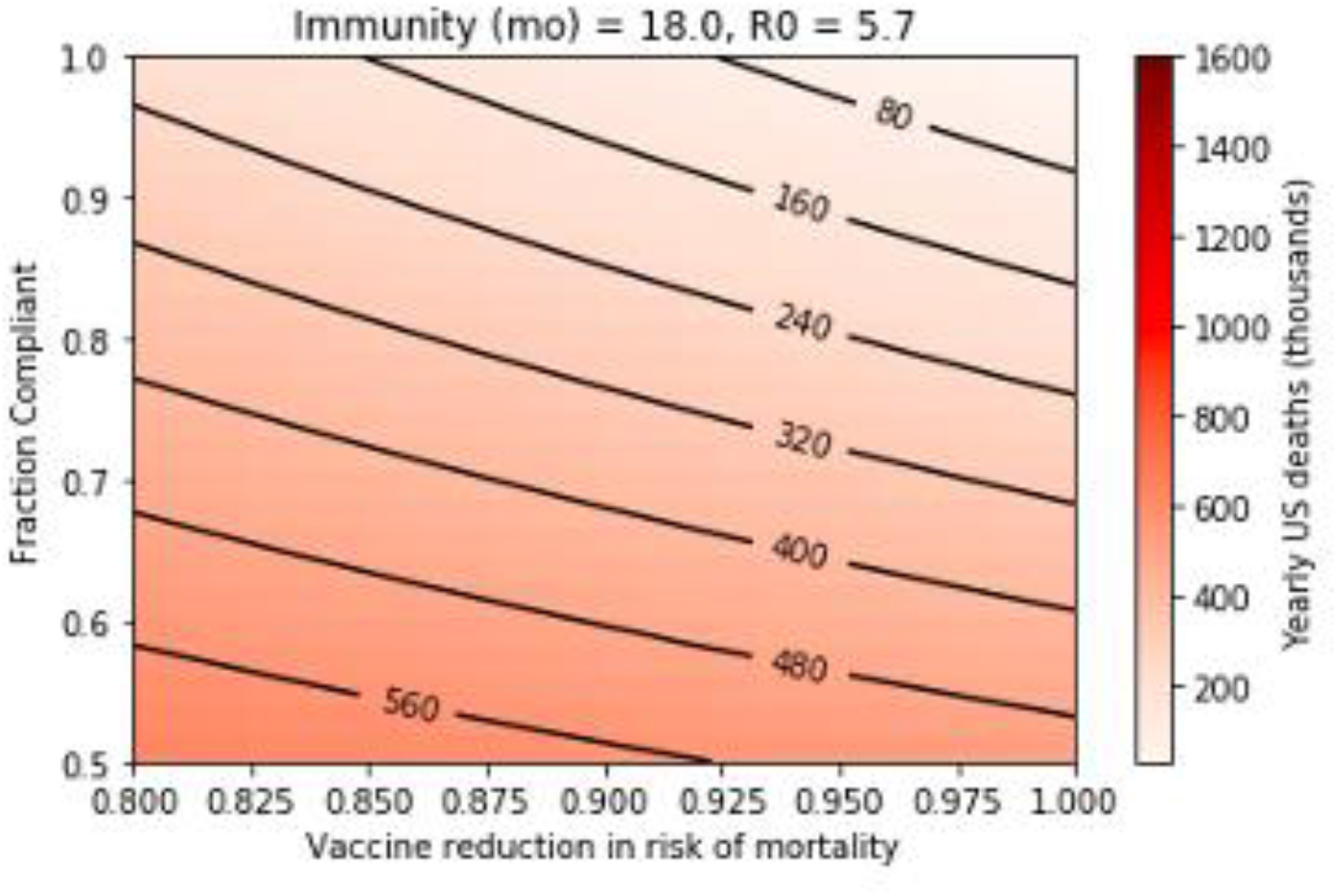
Steady-state yearly US COVID-19 fatalities after rollout of a vaccine that reduces risk of death (R_0_ of 5.7 with an 18-month duration of natural immunity).

**Figure 2.**
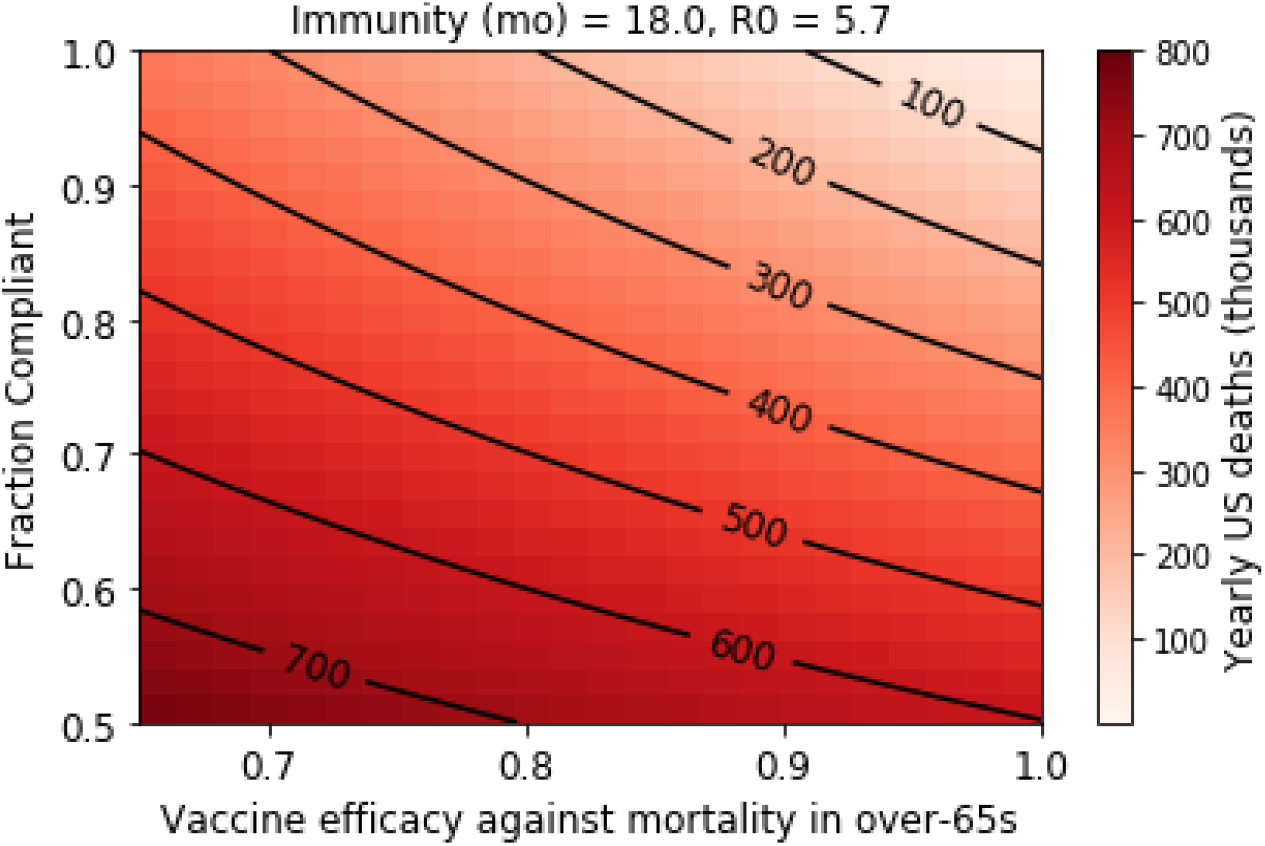
Range of mortality outcomes in the US population for a vaccine that reduces mortality risk by 95% in the under-65 population, and by varying degrees in the elderly. (Assumptions are: R_0_ = 5.7, 18-month immunity).

The impact of such a vaccine on COVID-19 deaths (Fig. 1) is explored. We note that in this parameter range, the duration of immunity appears to be a more significant determinant of yearly disease burden than R_0_ (Fig. S1). As expected, unless a perfectly effective vaccine is administered to all members of the population, symptomatic disease and COVID-19 deaths will persist indefinitely. Reduced rates of symptomatic disease and death can be achieved with an effective vaccine, but under pre-pandemic social and economic conditions, extremely high levels of vaccine effectiveness and population compliance are required to achieve “favorable” outcomes (for example, outcomes in which COVID-19 fatalities can be expected to be comparable to yearly influenza deaths.) If a vaccine with 97.5% efficacy against mortality is administered to 95% of the US population, approximately 80,000 yearly COVID-19 deaths can be expected if the duration of natural immunity is 18 months (Fig. 1). Relaxations in compliance or vaccine efficacy from this highly optimistic case result in large mortality burdens. Deaths remain high as compliance approaches 100% in most scenarios; this means that vaccinated individuals remain at risk of negative health outcomes unless vaccine efficacy is perfect. These simulations assume a 40% reduced rate of transmission in the vaccinated population; figures S7 and S8 demonstrate that this level of transmission reduction has a minimal impact on the yearly death toll.

### Reduced efficacy in the 65-and-older population presents a serious mortality risk

We conducted a sensitivity analysis on vaccine efficacy in the over-65 population in order to better understand the impact of age-dependent efficacy. For example, a vaccine that is 95% effective in all age groups and administered to 90% of the population can be expected to control US COVID-19 fatalities to the level of approximately 160,000 yearly (Fig. 1). If this vaccine were 95% effective in Americans younger than 65 but only 85% effective in older vaccinees, the model-predicted yearly death toll doubles to approximately 300,000 (Fig. 2). Clearly, vaccine efficacy in the over-65 age group is a highly important determinant of yearly COVID-19 mortality, and uncertainty in vaccine efficacy in this vulnerable population must be addressed.

### Elimination of SARS-CoV-2 is possible with a vaccine that is highly effective at preventing infection and widely administered

Elimination of SARS-CoV-2 is attainable with a vaccine that is highly effective against SARS-CoV-2 infection and with a high degree of vaccine compliance (Fig. 3). Elimination results from virtually zero disease transmission at steady-state because vaccinal immunity in the population impedes sustainable spread of the virus. The value of R_0_ defines the combinations of compliance and vaccine efficacy that result in successful elimination, while the duration of natural immunity determines the yearly disease burden if elimination is not achieved (Fig. S1).

**Figure 3.**
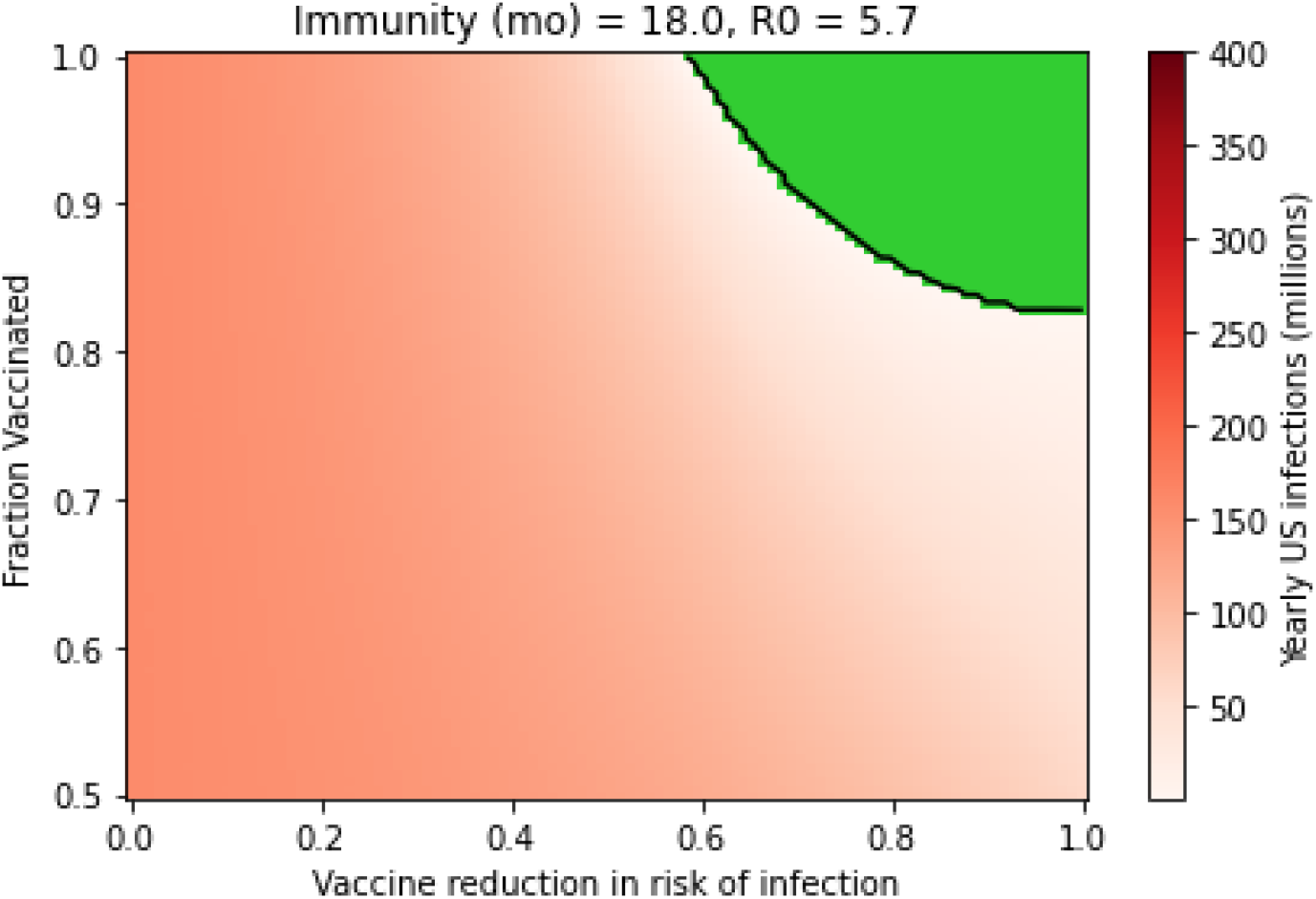
Elimination of SARS-CoV-2 can be achieved with a highly effective vaccine that is administered to most of the population. Green region represents cases where elimination is achieved, with virtually zero infections yearly. (Assumptions are: R_0_ = 5.7, 18-month immunity).

At an R_0_ of 5.7, at least 60% efficacy against infection is required (Fig. 3), while at an R_0_ of 3.32, 45% efficacy is required (Fig. S1); in both of these scenarios, for this level of efficacy against infection to suffice, perfect compliance with vaccination is required. For a vaccine that prevents infection, deviations from perfect compliance can be compensated by improvements in vaccine efficacy, but even if the vaccine is perfectly effective, a minimum of 82% compliance is required to achieve COVID-19 elimination under an R_0_ of 5.7, and 70% compliance is required if the R_0_ is 3.32. Elimination of SARS-CoV-2 with an infection-preventive vaccine represents a stable strategy: a variety of compliance conditions and vaccine efficacies are acceptable, and if vaccinal immunity in the population exceeds the threshold for elimination, small changes in vaccine efficacy or compliance can be tolerated without impacting disease spread.

### The vaccine’s effect on transmission by vaccinated infected individuals impacts potential for elimination

Clinical trials generally describe the protective efficacy of a vaccine – that is, the likelihood of illness in a vaccinated individual relative to an unvaccinated individual – but rarely measure the difference in likelihood of transmission for an infectious vaccinated individual relative to an infectious unvaccinated individual [58]. In Figure 3, we optimistically assumed that the vaccine’s efficacy against transmission in vaccinated infected individuals is equal to its efficacy against infection. However, it is possible that any SARS-CoV-2 vaccine exerts a more limited or nonexistent reduction in the likelihood of onward transmission. Figure 4 mirrors Figure 3 while assuming the vaccine has no impact on the likelihood of transmission in infected vaccinated individuals. This change significantly shrinks the available solution space, meaning that a higher degree of compliance and especially vaccine efficacy against infection is required to achieve elimination of SARS-CoV-2: In this case, at least 82% efficacy against infection is necessary if all members of the population are vaccinated and the R_0_ is 5.7.

**Figure 4.**
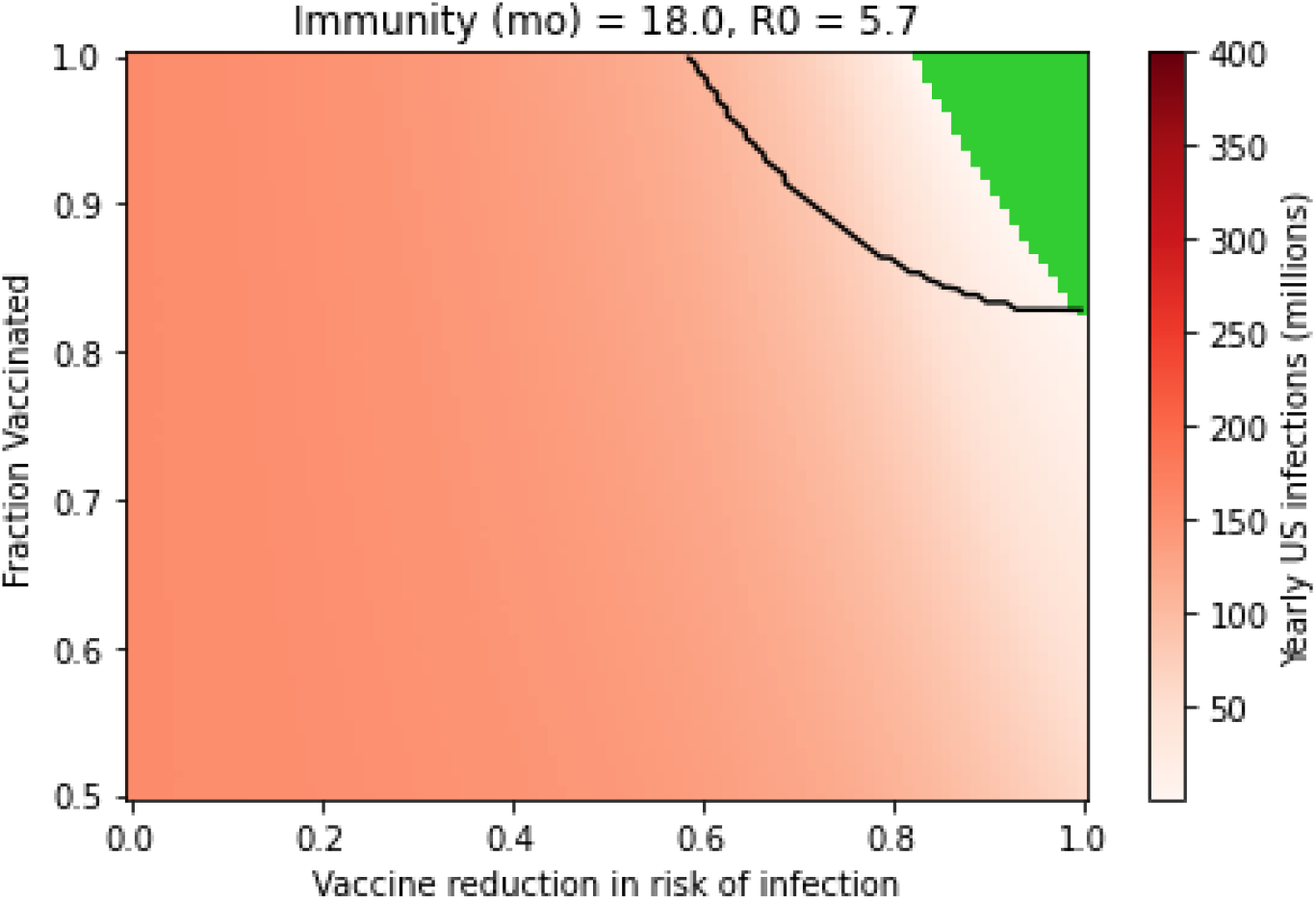
A vaccine that prevents infection but not transmission requires a higher degree of efficacy and compliance to achieve elimination. Black lines outline the elimination space for a vaccine that prevents infection and transmission to an equal degree, as shown in Figure 3. Green region represents cases where elimination under the vaccine is achieved, with virtually zero infections yearly. (Assumptions are: R_0_ = 5.7, 18-month immunity).

Figure S6 further demonstrates the importance of the vaccine’s impact on onward transmission as an aspect of disease elimination strategy: the extent to which a vaccine reduces transmission can determine whether the vaccination campaign is successful in eliminating disease. In fact, from a public health perspective, the vaccine’s efficacy against onward transmission is as impactful on population disease burden as its efficacy against infection.

### Complementary interventions improve potential for SARS-CoV-2 elimination

Interventions that can be stacked on top of a vaccine, such as masking and antiviral or passive immunity-based prophylaxis, can be deployed to reduce the thresholds for vaccine efficacy and compliance required for SARS-CoV-2 elimination (Fig. 5). These measures provide a valuable hedge against imperfect compliance with a vaccine, heterogeneity in regional transmission dynamics, limited vaccinal protective efficacy, waning vaccinal immunity, and unknown vaccinal impact on transmission. Complementary interventions improve the accessibility of elimination across a wide range of possible efficacies and compliance levels. A highly effective (95% [59]) intranasal prophylactic used by 50% of the population, for example, substantially reduces both the compliance and the vaccinal efficacy required to achieve elimination. Universal masking, which can be expected to reduce risk of infection by at least 50% [60], remains a highly effective measure but represents a significant divergence from pre-pandemic activity (the “new normal” as opposed to the “old normal”).

**Figure 5.**
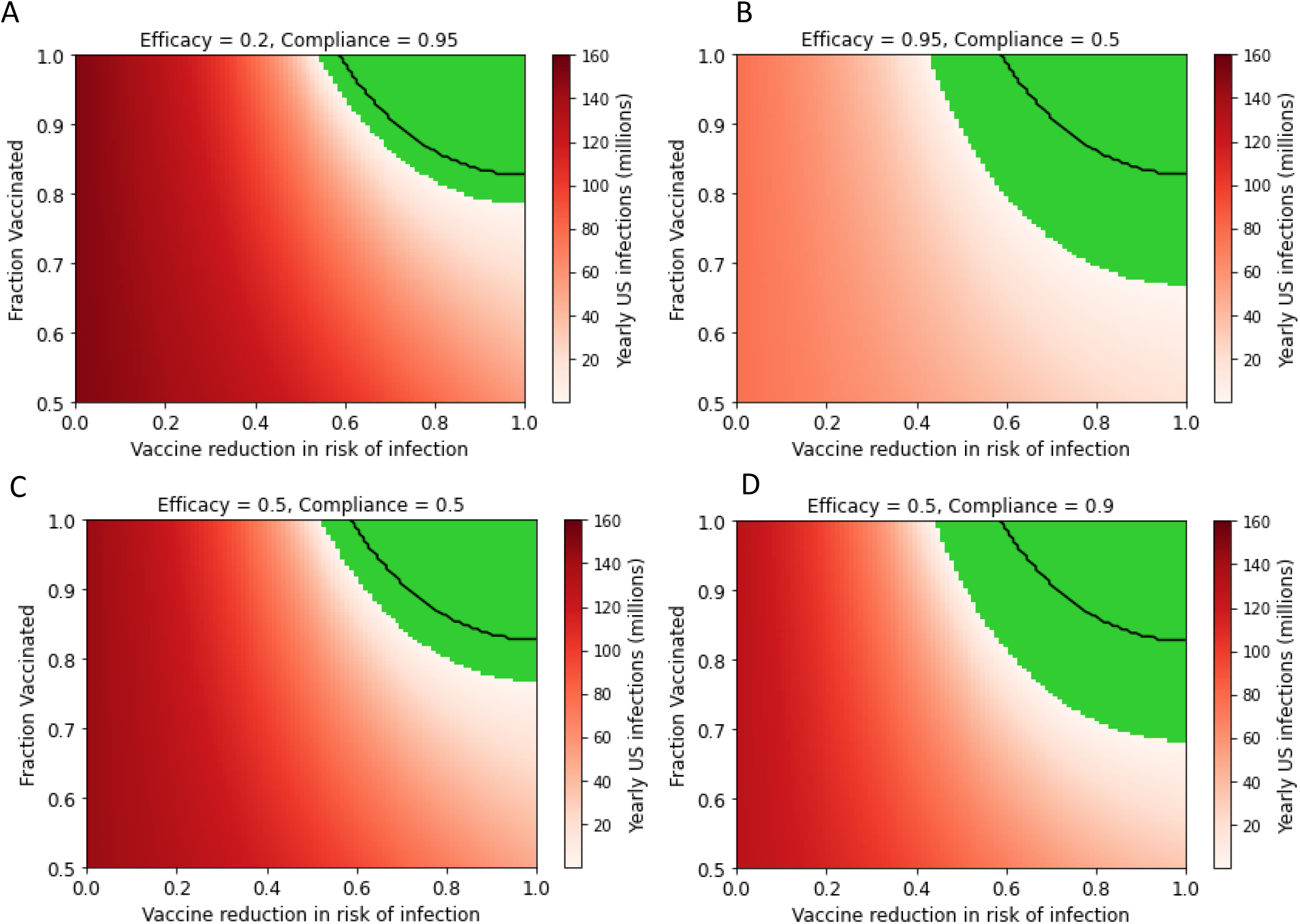
Interventions complementary to the vaccine improve elimination potential of a vaccine (Assumptions are: R_0_ = 5.7, 18-month immunity). Black lines outline the elimination space for the vaccine alone. In the figure panels, four example complementary interventions are explored: A) a mutually compatible, competing vaccine achieving 50% reduction in risk of infection and 50% compliance in the population; B) universal masking, which reduces the risk of infection by 50% and reaches 90% compliance; C) a passive intervention, such as improving ventilation in schools and workplaces, which impacts 95% of the population but has a small 20% impact on risk of infection; and D) a highly effective (95%) intranasal prophylactic that 50% of the population uses.

## Discussion

In this work, we sought to understand the role that the first wave of SARS-CoV-2 vaccines would play in bringing about a return to normalcy. By definition, a return to normalcy implies a return to pre-pandemic levels of contact, and with this in mind have used an R_0_ that corresponds to published estimates from the early stages of the pandemic [46]. Our goal in this work was not to predict the future broadly, but rather to focus narrowly on potential limitations of the first wave of vaccines, and ask “what if” questions around the epidemiological consequences of these potential limitations.

A significant potential limitation of the first wave of SARS-CoV-2 vaccines is the failure to sufficiently prevent infection and transmission in the vaccinated population. While this has not been demonstrated conclusively, it remains a plausible risk that has been observed for other vaccines in the past and that is not being addressed directly in the vast majority of ongoing clinical vaccine studies [6,56]. In this study, we have used mathematical modeling to understand conditions in the United States at steady state, if a vaccine that was unable to prevent infection and exerted limited reduction in transmission was used as the primary or sole public-health intervention. Our results paint a grim picture: at steady state, we could expect 30 million symptomatic COVID-19 cases a year for a vaccine that is 95% effective at reducing symptoms and that is taken by 80% of the population (using best-estimate assumptions of natural immunity that lasts for 18 months, permanent vaccine immunity and an R_0_ of 5.7). If the reduction in risk of symptoms was concomitant with a reduction in risk of mortality (a best-case assumption), this scenario would lead to 240,000 deaths per year, making COVID-19 the third leading cause of death in the United States [51].

Another significant potential limitation of the first wave of SARS-CoV-2 vaccines is reduced protection for the elderly. This remains a plausible risk as of this writing and would be consistent with the behavior of vaccines for a number of other diseases [30,32]. We used our modeling framework to explore the impact of a vaccine that was less effective in the elderly. Again, the picture is grim. For example, for a vaccine that is 85% effective in the over-65 population, at steady state, we could expect around 400,000 deaths a year (using the assumptions of 95% vaccine efficacy in the under-65 population, 80% compliance, 18 months natural immunity, permanent vaccine immunity and an R_0_ of 5.7; Fig. 2). The vast majority of these deaths would occur in the over-65 population. Crucially, for the over-65 population, a vaccine that does not prevent transmission and that is only partially effective will result in a never-ending pandemic. Beyond the scope of this paper, there are clear implications for the age structure of the US population and life expectancy in general.

The work described here has a number of limitations. First, we assumed that the populations are well-mixed, and did not include stochastic effects. As the focus in this work is examining steady-state mortality and morbidity burdens, this choice is justified as stochastic effects are unlikely to dominate steady-state behavior in a situation with widespread endemic transmission of SARS-CoV-2. Second, we assumed that vaccinal immunity was permanent. The available data for the front-runner vaccines is not consistent with this assumption [5], and waning vaccinal immunity will compound the limitations of real-world vaccine effectiveness described here, further eroding the public-health impact of vaccines that do not prevent transmission. A further limitation of our work is that we focused on the steady-state conditions for the pandemic. As a result, our work does not (seek to) predict the near-term kinetics of COVID-19 within the United States over the next year. We also focused the work narrowly on the United States – while the modeling is generalizable, our mortality and morbidity estimates were focused on the numbers for the US, to make the argument easier to understand. We assumed that stacking interventions (Fig. 5) were adopted independently by individuals. A full exploration of the impact of individual choices on the trajectory of the pandemic is beyond the scope of this paper, but has been explored thoroughly by us and others [61,62]. Lastly, the availability of vaccinal efficacy data limits our analysis. To address this uncertainty, we have performed sweeps to demonstrate the sensitivity to efficacy parameters. We have assumed, however, that the vaccines’ efficacy against mortality will be at least 80%; however, there is insufficient data in the current publications to determine the impact of the vaccines on mortality (in both the Pfizer and Moderna studies, only one COVID-19-related death was reported [8,56]).

Under the scenarios examined in this paper, SARS-CoV-2 will become endemic within the population. This outcome has been discussed in the popular press, and is often referred to as “learning to live with the disease” [63,64]. This outcome is in contrast to the “go for zero”[65] outcome focused on disease elimination, being pursued by countries such as Australia, New Zealand, China, Taiwan and Korea. Our work shows what “learning to live with the disease” would look like. For the young and healthy, once vaccinated, there may be little incentive to comply with further public health measures aimed at limiting the spread of SARS-CoV-2. If mask-wearing was difficult to incentivize in the population before the emergence of a vaccine, it stands to reason that vaccination will make it more difficult, not easier [62]. For those with limited access to the vaccine or with limited benefit from it, the pandemic will never end.

At a national level, “learning to live with the virus” is a fragile solution that would make the country critically dependent on the vaccine supply chain and vulnerable from a national security perspective to actions by both state and non-state actors. The extensive logistical planning and infrastructure required for maintenance of population vaccinal protection from COVID-19 morbidity and mortality could be readily disrupted by natural disasters, domestic unrest, foreign interference, or future pandemics, resulting in the emergence of symptomatic and deadly COVID-19 in the wake of unrelated crises.

Beyond potential human adversaries, the virus itself is a formidable opponent. In other work [10], we have demonstrated the potential risk of evolutionary immune evasion that comes with strategies that focus on the viral spike protein. Natural selection can be expected to favor increased transmissibility [66], and a transition to increased transmissibility has been shown to occur even after pandemic onset for H1N1 [67,68], and for SARS-CoV-2 during the current global crisis [69,70]. Farming SARS-CoV-2 virus at scale with a vaccine that fails to mitigate transmission is an invitation to disaster on this front, as viral evolution over time will limit the space within which a vaccine-mediated strategy can be effective. More transmissible and possibly antibody drug-evading variants of SARS-CoV-2 have already emerged, even before the widespread deployment of antibodies and vaccines [71,72].

If the choice is made to pursue SARS-CoV-2 elimination, however, existing vaccines can still play a very positive role. The current crop of vaccines represents one more layer in the “Swiss cheese strategy” [73–75], as long as it is made clear to vaccinated individuals that they (counterintuitively) may face increased risk after being vaccinated and returning to pre-pandemic lifestyles. To use an analogy, wearing a seatbelt reduces risk of death when driving, but should not be taken as license to down three beers before taking the wheel. Public health messaging should emphasize that the benefit from the vaccine can be offset by actions taken by individuals to increase their contact rate, so compliance with public health measures to reduce transmission will remain in the individual’s best interest. Crucially, public health strategy should diversify its approach, broadening focus to passive interventions such as ventilation and complementary biomedical interventions such as intranasal prophylactics. These interventions can be stacked on top of a successful vaccine program to increase the likelihood of eliminating SARS-CoV-2 and to address risks of evolutionary escape, reduced protection in some subpopulations, vaccine noncompliance, and limited efficacy against transmission. Further study of the current vaccines’ performance characteristics (impact on infection/ transmission, efficacy in older vaccinees, durability of protection, impact on mortality and morbidity) will be required to better understand their public health impact.

Although it is possible the current crop of vaccines provide sufficient protection from infection and transmission to achieve elimination, there are insufficient data to support this currently. There is much discussion regarding the “herd immunity threshold” and the corresponding fraction of the population that must be vaccinated to achieve herd immunity [76]. These discussions often neglect the fact that the extent of population immunity is determined by both the fraction of individuals vaccinated and the effectiveness of the vaccine against transmission and infection. There is a minimum threshold of the population requiring vaccination to achieve herd immunity with a vaccine that provides perfect sterilizing immunity, and correspondingly there is a minimum threshold for vaccine efficacy against infection and transmission to reach herd immunity if the entire population is vaccinated. Perfect protection from infection and transmission is not consistent with the preclinical data for these vaccines [25], the frequency of asymptomatic carriers in the AstraZeneca trial [6], or the theorized “maybe 50%” reduction in transmission described by Moderna and Pfizer [77,78].

Notably, this analysis covers a subset of the unhedged risks involved in managing the COVID-19 crisis with existing vaccines: age-dependent efficacy, poor vaccine uptake in the population, and unknown efficacy against transmission and infection. The risks of evolved resistance to vaccines, of selection for increasingly transmissible strains of SARS-CoV-2, of waning vaccinal immunity, and of logistical challenges in the induction and maintenance of population-level vaccinal immunity are beyond the scope of this work but layer on top of the risks evaluated in this work.

Our work also shows that there is a path forward for vaccines to contribute to SARS-CoV-2 elimination. For vaccines that stop infection and/or transmission, these can be leveraged to eliminate the disease with even moderately high levels of compliance, a point that has been made by others as well [79]. We submit that the rate of infection and viral load (a predictor of transmissibility [80]) are critical public-health metrics for successful elimination and may be more meaningful than the number of symptomatic cases. Crucially, stacking interventions (such as indoor air filtration and prophylactics) can help broaden the range of vaccine efficacy and compliance that leads to successful elimination. Complementary vaccines (double vaccinations) may also represent a feasible path forward.

The race to develop vaccines for COVID-19 has yielded an unprecedented technological achievement for mankind, one that would have been inconceivable a decade ago. Martial analogies abound, with the “Manhattan project”-like efficiency of Operation Warp Speed [81], yielding the “weapon that will end the war” [82] Our work suggests a different military analogy: the bombard cannons of the Hundred Years War promised more firepower than they delivered, and often blew up in the faces of their operators [83–85]. Even so, commanders with a precise understanding of the limitations of these early cannons were able to incorporate them into combined-arms tactics that yielded victories on the battlefield. Clear-eyed rationality about the limitations of these early weapons also allowed for their systematic optimization, and the bombard cannons of the 14^th^ century were iteratively improved until they met their promise, and matured into game-changing weapons.

## Data Availability

Please contact AC for simulation code.

