## Supplemental figures for "Beyond the new normal: assessing the feasibility of vaccine-based elimination of SARS-CoV-2"

### Supplemental Methods and Results

*Herd immunity strategy results in extensive endemic disease*

Fringe groups have proposed allowing COVID-19 to spread freely among the general population with the hope that natural immunity will eventually slow the spread of disease. Although the SEIRS model demonstrates that a strategy entirely dependent on natural immunity would eventually lead to stabilization of SARS-CoV-2 transmission, the steady-state that is reached involves catastrophic disease burden and mortality (Figure S7). Although estimates of the $R_0$ and duration of immunity of SARS-CoV-2 vary, optimistic estimates for these parameters still result in more than 100,000 COVID-19 deaths annually. Under no relevant conditions does the pandemic extinguish itself without intervention. Our best estimate for $R_0$ is 5.7 based on CDC data and for the duration of immunity is 18 months based on the time-to-baseline for anti-SARS-CoV-2 antibodies post-infection. Under these conditions, the model predicts 176 million annual US infections and 1.2 million annual US deaths if no action is taken to contain SARS-CoV-2.

### Supplemental Figures

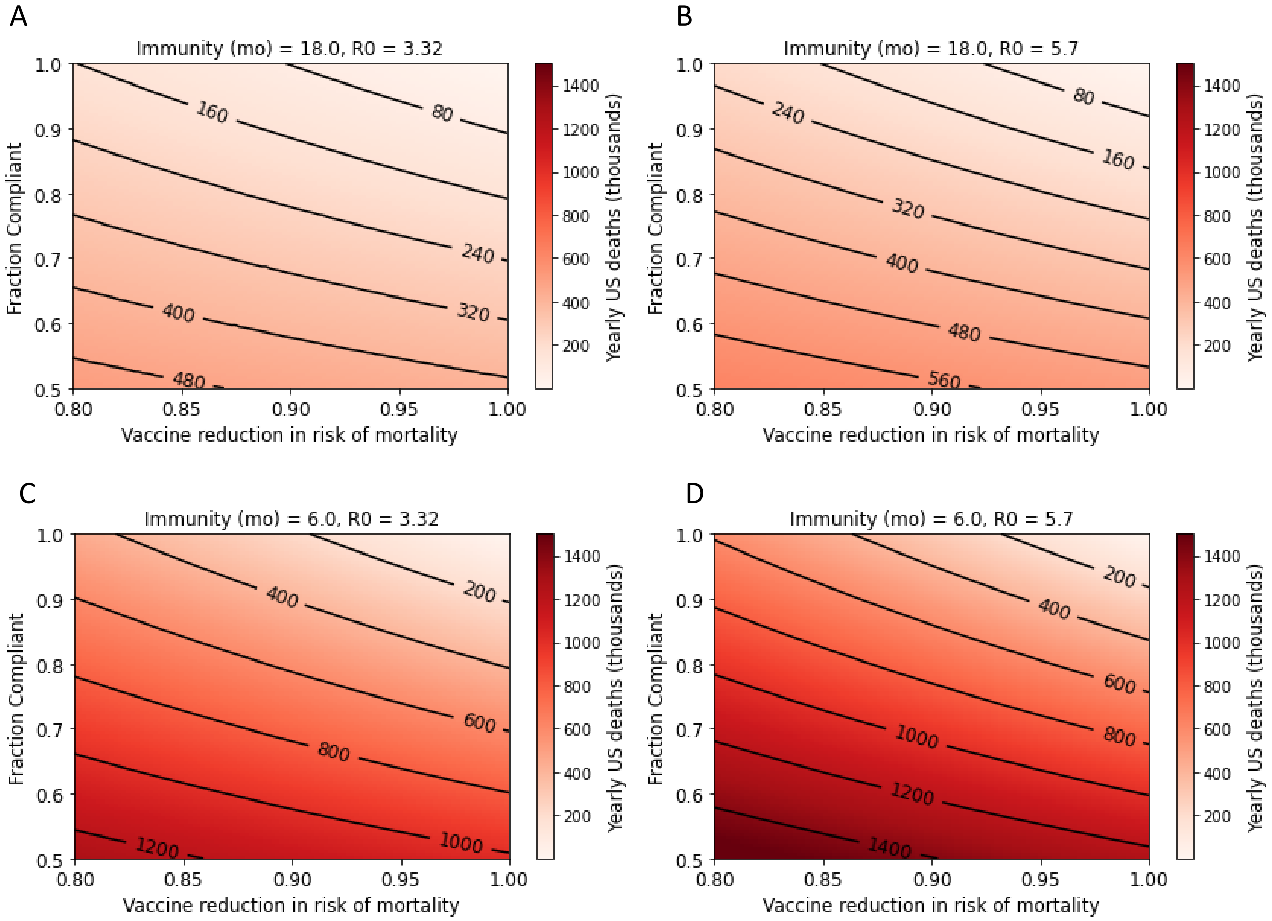


**Figure S1.** Steady-state yearly US COVID-19 fatalities after rollout of a vaccine that reduces risk of death. This figure is parallel to Figure 1 in the main text but explores four sets of parameters for the duration of natural immunity and $R_0$.

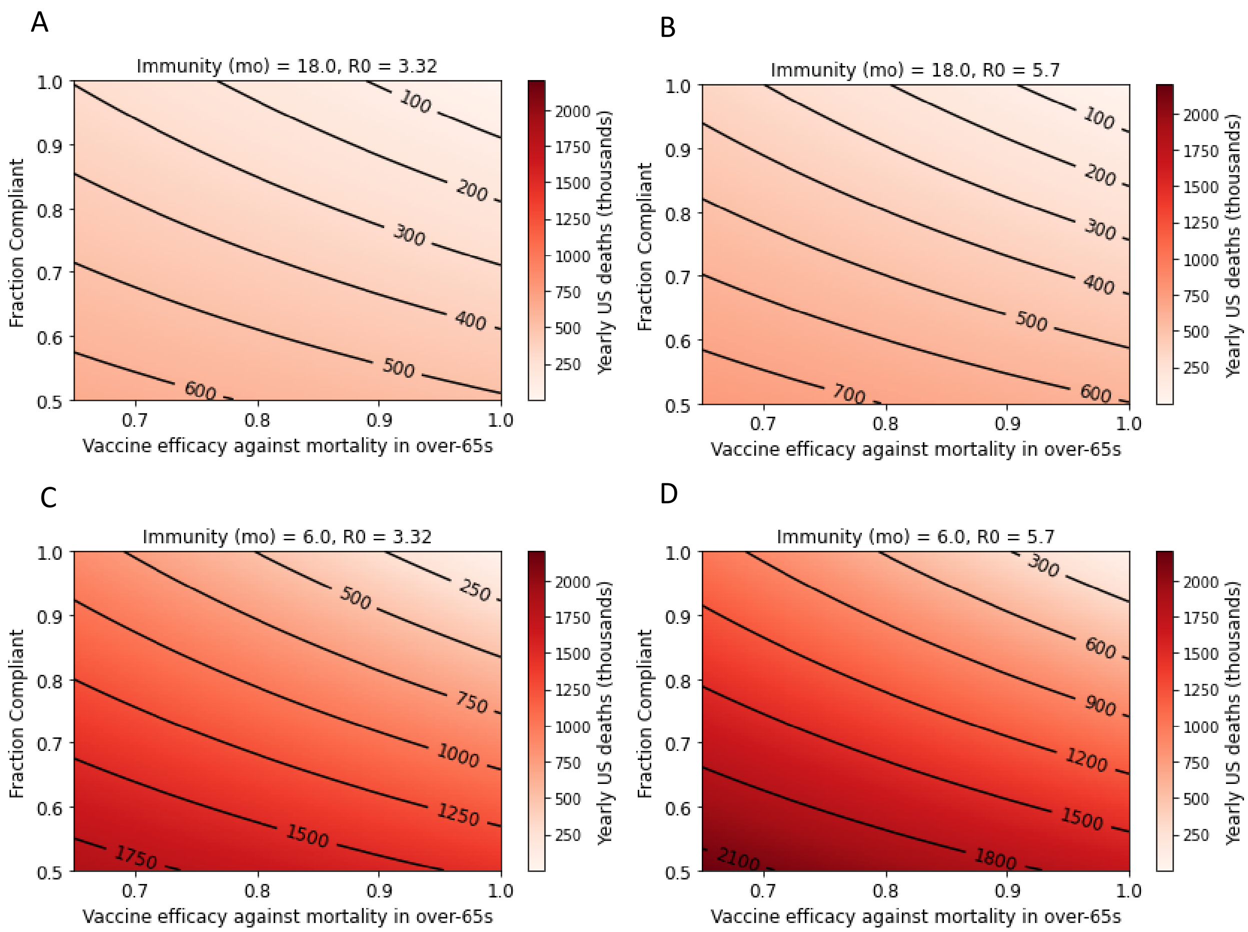


**Figure S2.** Steady-state US yearly deaths after deployment of a vaccine that reduces mortality risk by 95% in the under-65 population and by varying degrees in the elderly. This figure is parallel to Figure 2 in the main text but explores four sets of parameters for the duration of natural immunity and $R_0$.

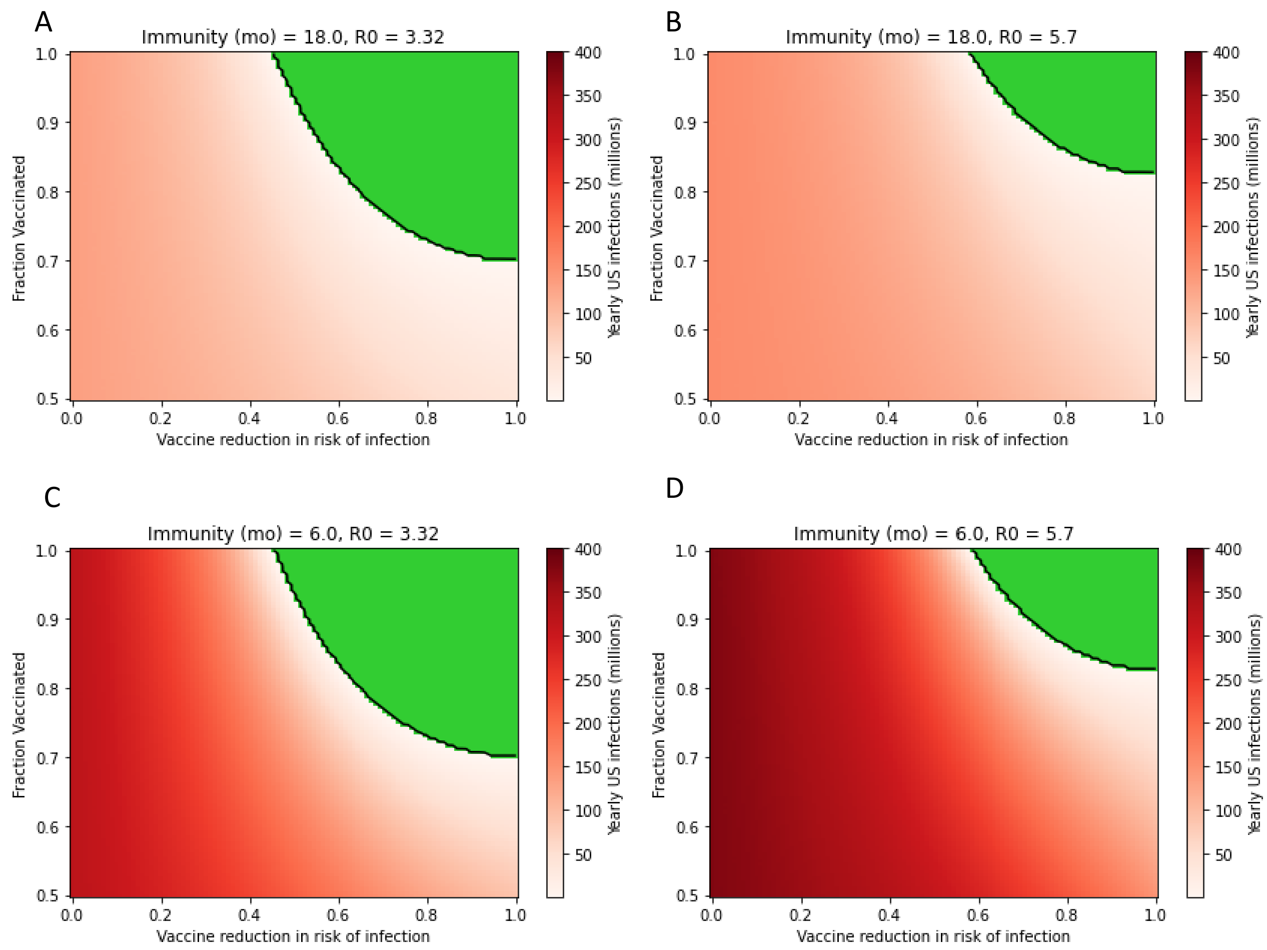


**Figure S3.** Steady-state US yearly SARS-CoV-2 infections after deployment of a vaccine that reduces risk of infection and transmission. This figure is parallel to Figure 3 in the main text but explores four sets of parameters for the duration of natural immunity and $R_0$. Green region represents regime in which SARS-CoV-2 is eliminated in the population and yearly infections approach zero.

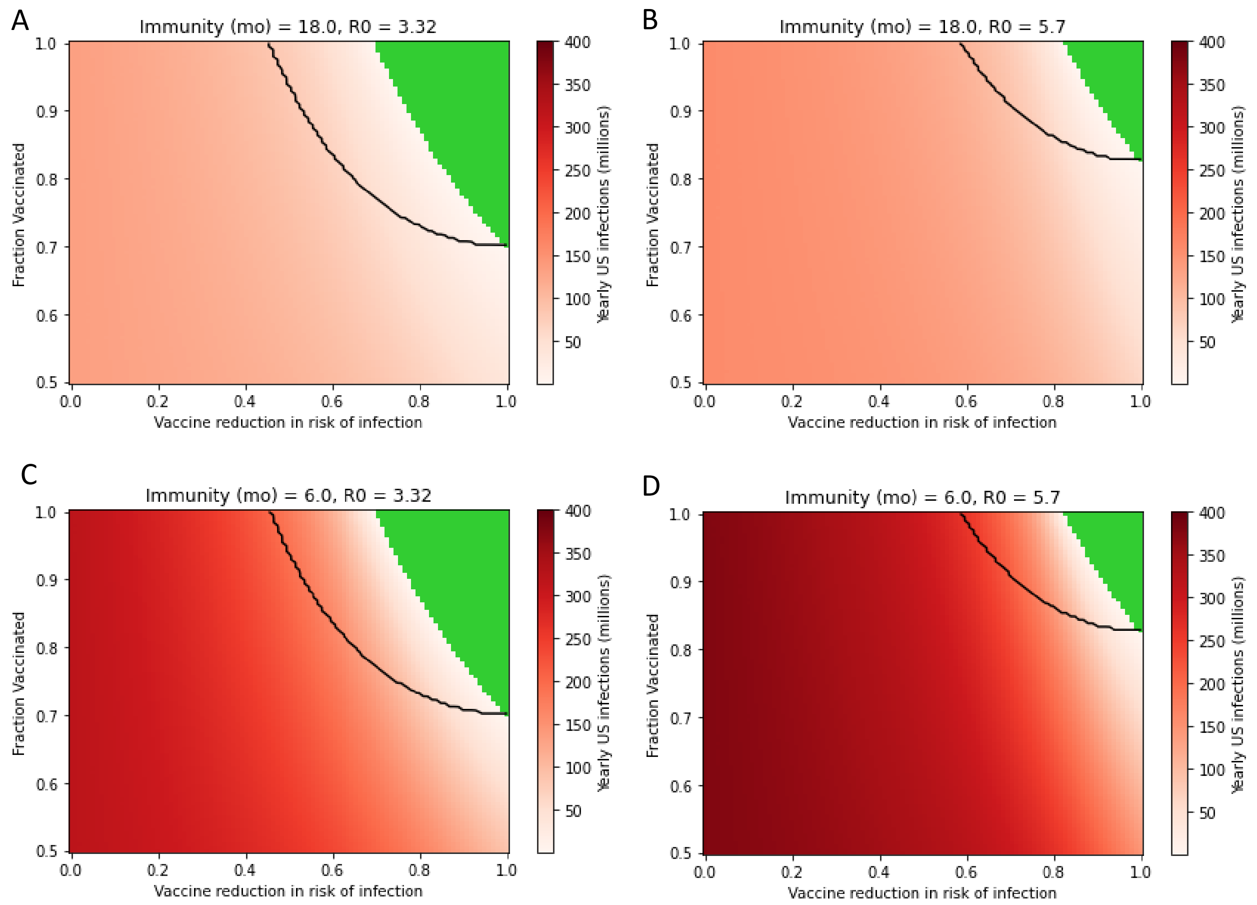


**Figure S4.** Steady-state US yearly SARS-CoV-2 infections after deployment of a vaccine that reduces risk of infection but not transmission. This figure is parallel to Figure 4 in the main text but explores four sets of parameters for the duration of natural immunity and $R_0$. Black lines outline the elimination space for a vaccine that prevents infection and transmission to an equal degree, as shown in Figure S3. Green region represents cases where elimination under this vaccine is achieved, with virtually zero yearly infections.

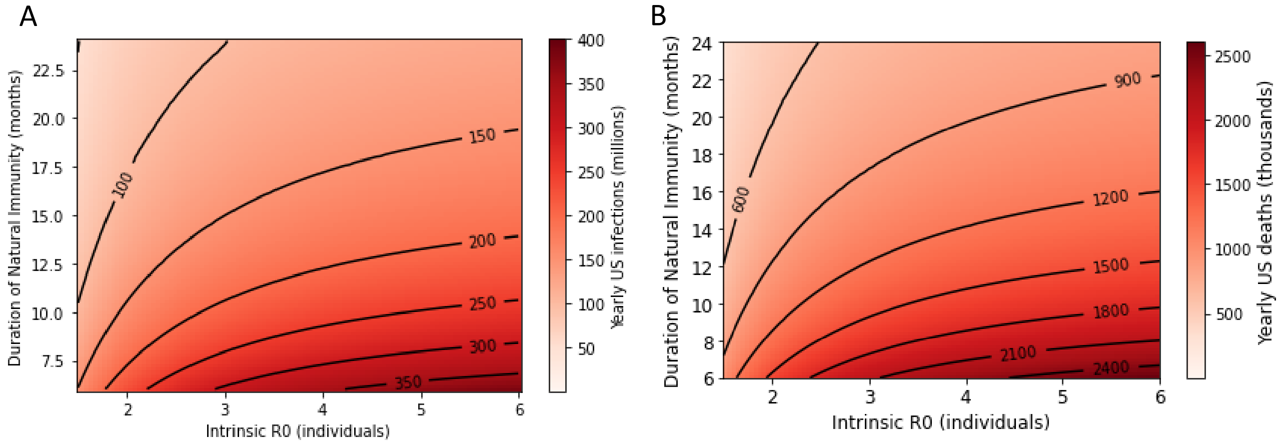


**Figure S5.** Natural herd immunity is no path to normalcy. US yearly infections (A) and COVID-19 deaths (B) are predicted at steady-state for a variety of $R_0$ and duration of natural immunity estimates. Extensive disease and mortality burdens are expected under all endemic scenarios.

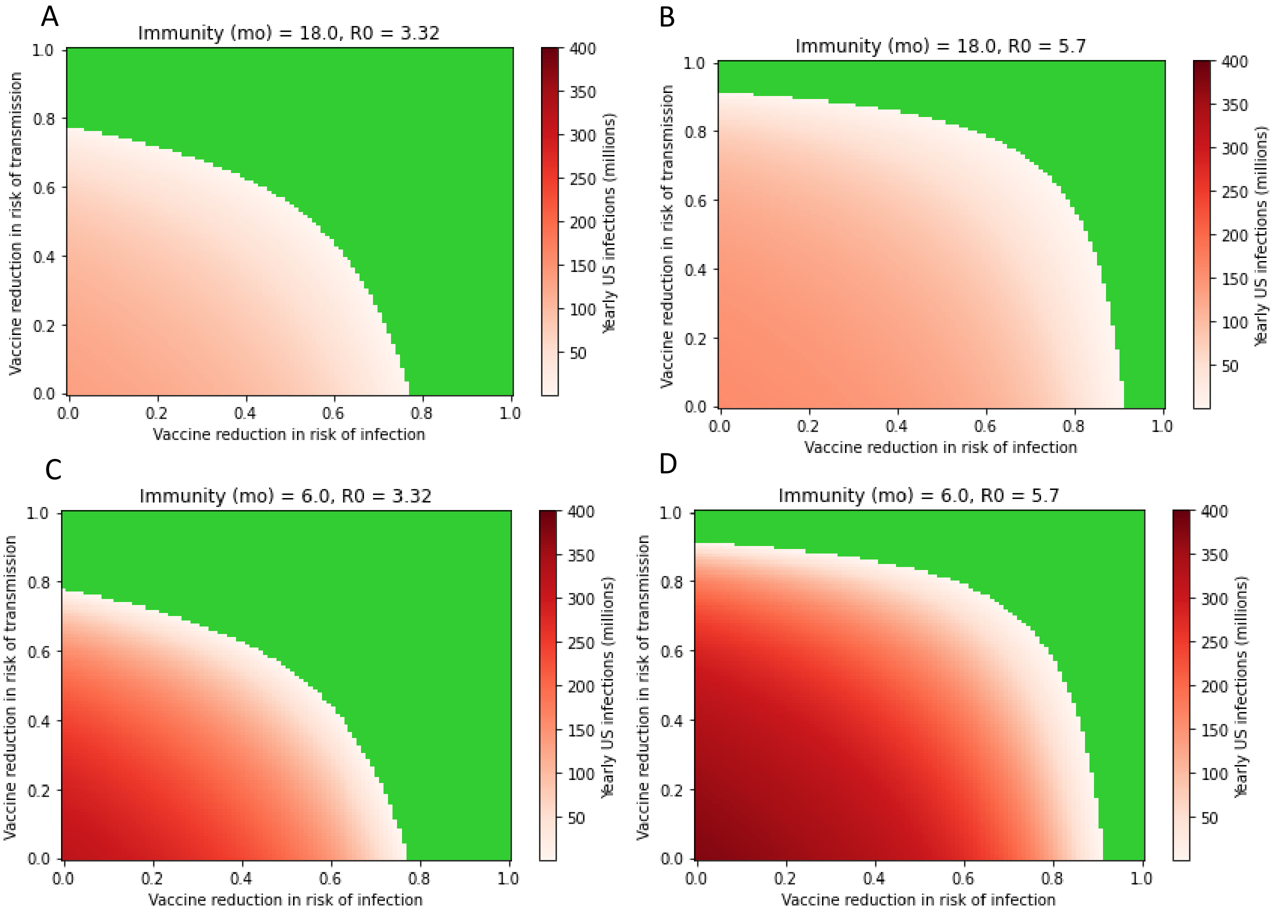


**Figure S6.** Impact of vaccine reduction in risk of infection and transmission on yearly US infections given 90% vaccine uptake in the population. Vaccine efficacy against transmission is equally impactful compared to vaccine efficacy against infection and determines success or failure to achieve eradication in many cases. Panels represent four possible scenarios: A) $R_0$ of 4 with an 18-month duration of natural immunity, B) $R_0$ of 5.7 with an 18-month duration of immunity, C) $R_0$ of 4 with a 6-month duration of immunity, D) $R_0$ of 5.7 with a 6-month duration of immunity.

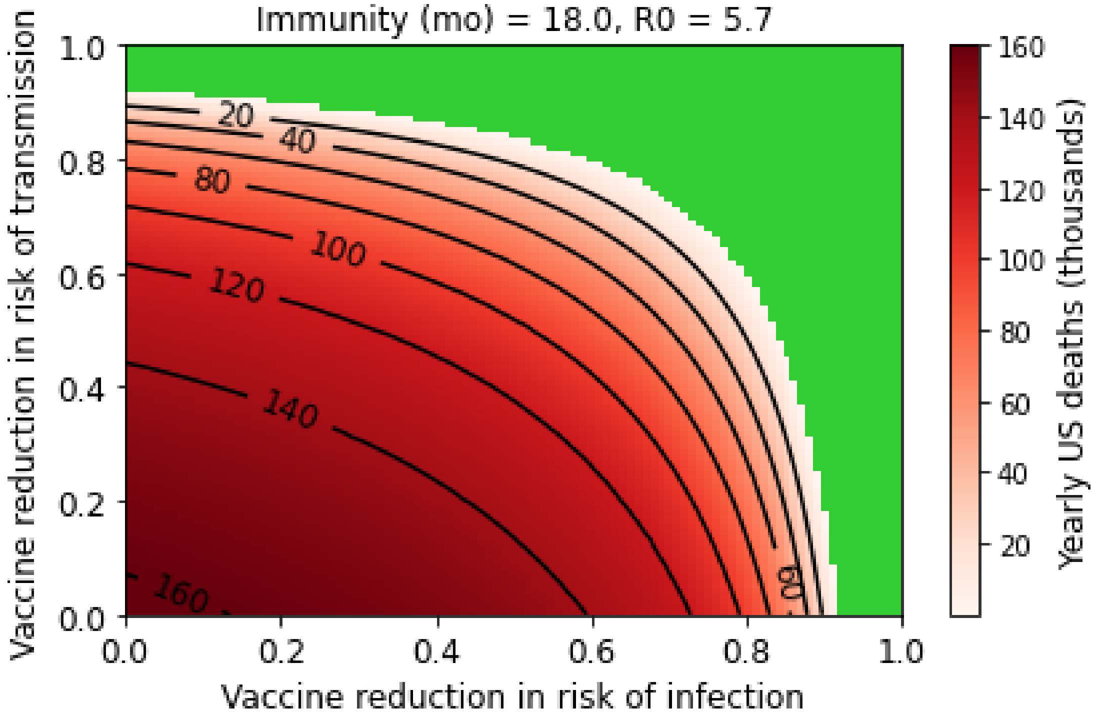


**Figure S7**. Relationship between vaccine efficacy against infection and transmission and US mortality. In this figure, 90% of Americans are assumed to be vaccinated and the vaccine is assumed to have an age-independent 95% efficacy against mortality.

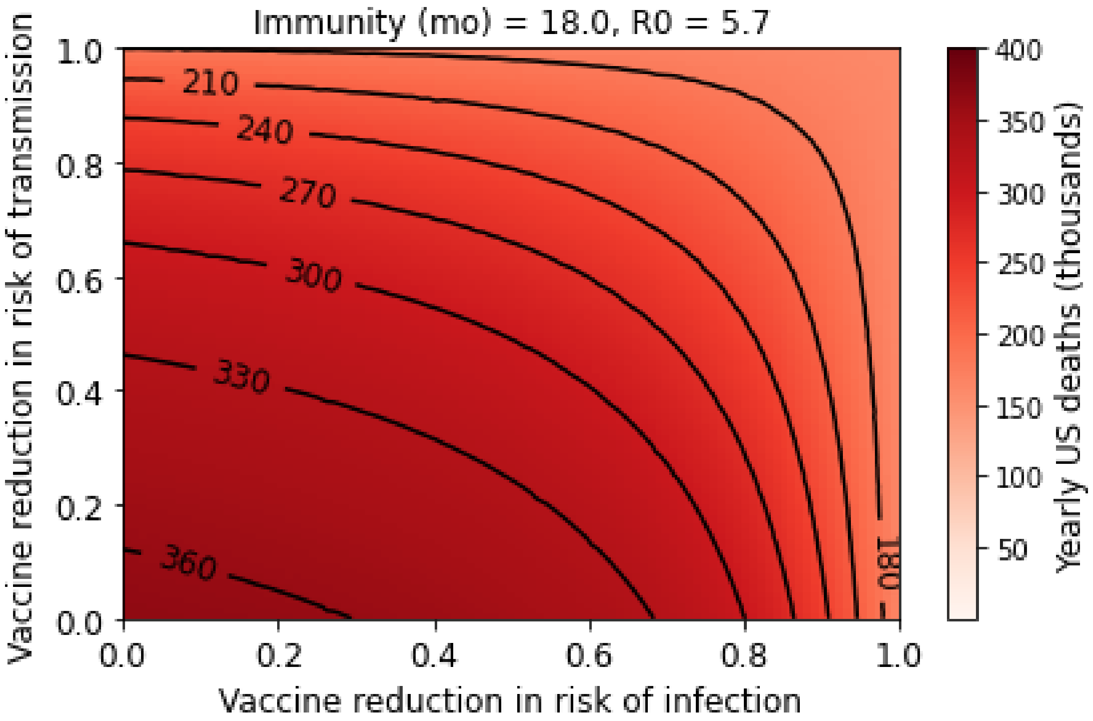


**Figure S8.** Relationship between vaccine efficacy against infection and transmission and US mortality. In this figure, 70% of Americans are assumed to be vaccinated and the vaccine is assumed to have an age-independent 95% efficacy against mortality. At this $R_0$, disease elimination is impossible with only 70% vaccine compliance.

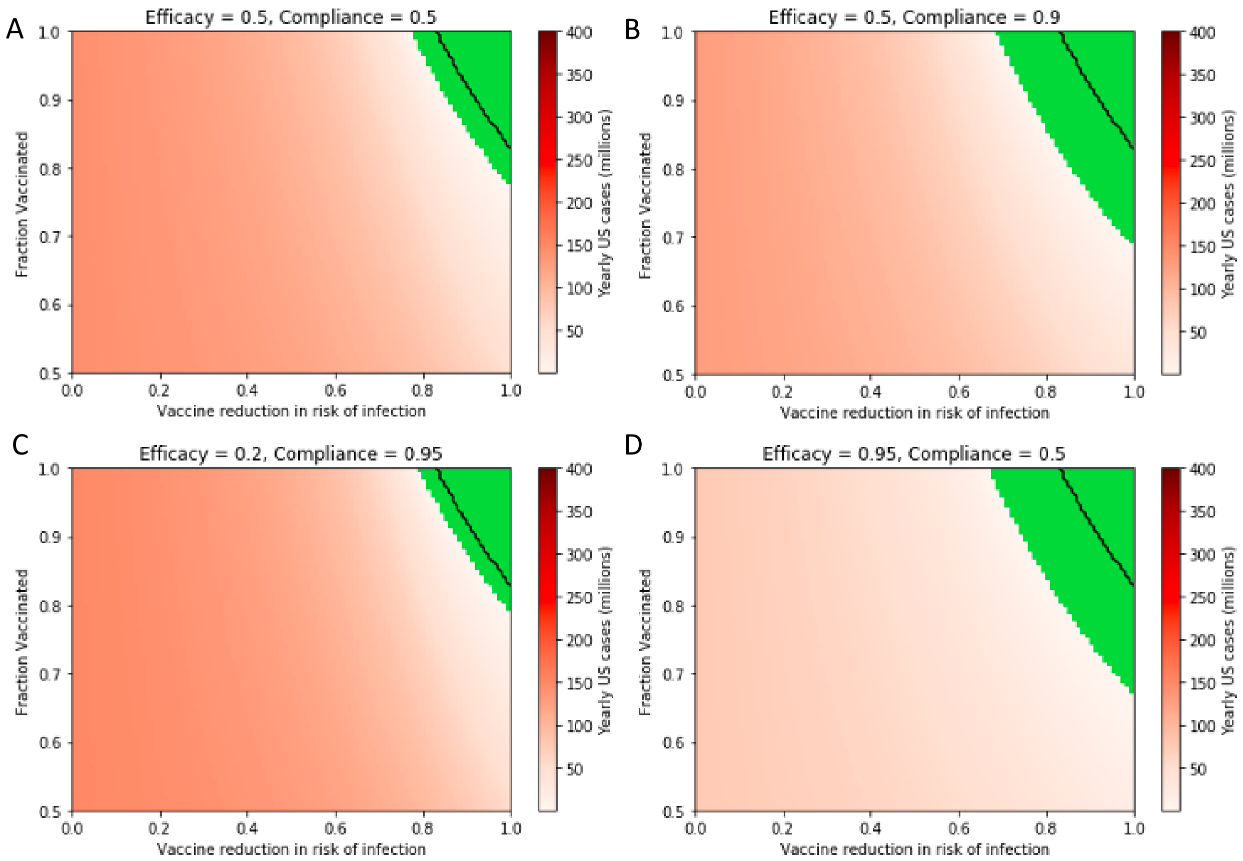


**Figure S9.** An eradication strategy involving a vaccine that prevents infection but not transmission is more likely to succeed if complementary interventions are in place. Black lines outline the eradication space for the vaccine alone. This figure is parallel to Figure 4; the vaccine is assumed to have no effect on transmission, the $R_0$ is assumed to be 5.7, and the duration of natural immunity is 18 months. Black lines outline the eradication space for the vaccine alone. In the figure panels, four example complementary interventions are explored: A) a compatible, competing vaccine achieving 50% reduction in risk of infection and 50% compliance in the population; B) universal masking, which reduces the risk of infection by 50% and reaches 90% compliance; C) a passive intervention, such as improving indoor ventilation, which impacts 95% of the population but has a small 20% impact on risk of infection; and D) a highly effective (95%) intranasal prophylactic that 50% of the population uses.

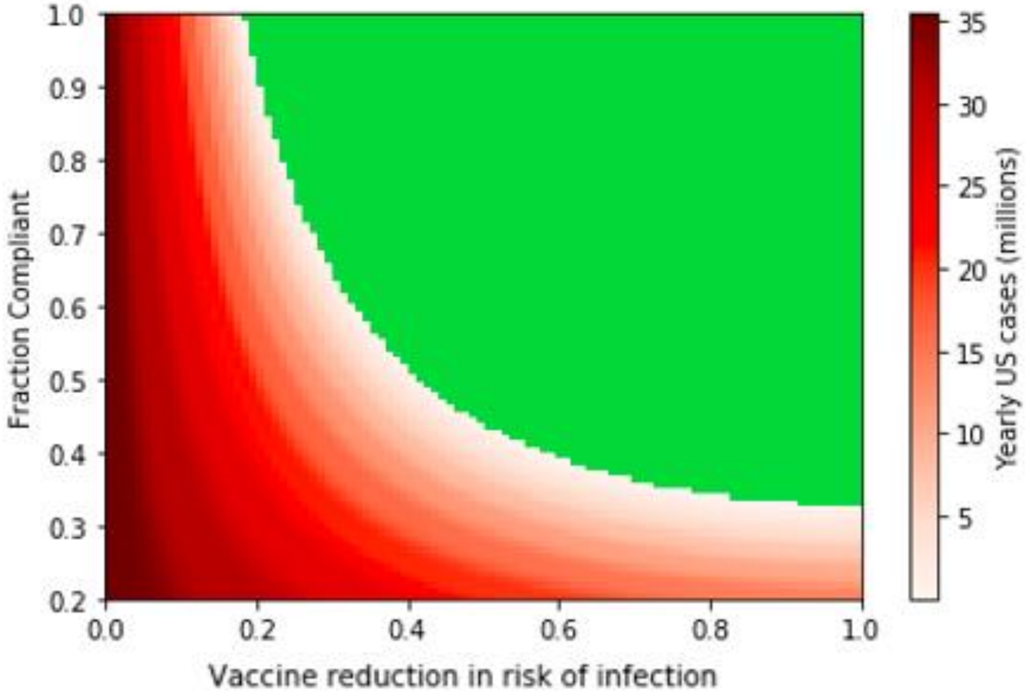


**Figure S10.** Influenza is eradicated more readily than SARS-CoV-2. Green region represents successful vaccine-based eradication based on an SEIR model for influenza [1].
